# The association between NfL and Cognitive Functioning in older adults diagnosed diabetes: a cross-sectional analysis from NHANES

**DOI:** 10.1101/2024.06.20.24309238

**Authors:** LiLi, Cheng-Bo Li, Li Zhu, Jiang Zhu

## Abstract

**Objective:** This research aimed to explore the correlation between serum Neurofilament Light Chain (NfL) levels and Cognitive Function in diabetic participants.

**Methods:** Utilizing cross-sectional data from the National Health and Nutrition Examination Survey (NHANES) 2013 to 2014, 173 individuals aged 60 years and above with diabetes were included in the analysis. Cognitive function was measured using the Animal Fluency exercise. Weighted multivariate linear regression analysis and a restricted cubic spline model were employed to assess the association between serum NfL levels and Animal Fluency Score (CFDAST) in diabetic patients.

**Results:** Serum NfL was categorized into weighted tertiles, with participants divided into groups Q1, Q2, and Q3. Significant differences in serum NfL were observed among the groups (P < 0.01). Participants in group Q3 exhibited higher levels of hemoglobin A1c and were older compared to groups Q1 and Q2 (P < 0.01). Furthermore, when serum NfL was treated as a continuous variable, it was found to have a negative correlation with CFDAST score (P < 0.01). Group Q1 had a higher CFDAST score than group Q3, as well as group Q2 (p for trend = 0.0021). A nonlinear relationship was observed between serum NfL and CFDAST score, displaying an inverted ‘U-shaped’ curve (P-overall = 0.01, P-for-non-linear = 0.0026).

**Conclusions:** Serum NfL has the potential to serve as a clinical biomarker for early detection of cognitive impairment in diabetic individuals aged over 60 years.

**Highlights:** Study about biomarkers for diabetes-associated cognitive decline is limited.

We found that there is an inverted U-shaped association between serum NfL and cognitive score.

Serum NfL may be a biomarker for early detection of cognitive decline in elderly diabetics.

## 1. Introduction

The co-occurrence of type 2 diabetes and cognitive dysfunction is prevalent worldwide(1), with type 2 diabetes being recognized as a significant risk factor for cognitive impairment and dementia(2). Growing epidemiological evidence supports a strong association between diabetes and cognitive impairment(3, 4). Individuals with diabetes experience a more rapid decline in cognitive functioning in the years following diagnosis(5). Cognitive deficits associated with diabetes, which represent the initial stage of cognitive decline in individuals with Type 2 diabetes, progress at a rate that surpasses approximately half of the normal cognitive aging process(6-10). It is crucial to identify early cognitive decline in patients with diabetes. However, the underlying mechanisms are still unclear, and current research on biomarkers for diabetes-associated impaired cognitive functions is limited and presents conflicting findings(11).NfL, an essential cytoskeletal component of large-diameter myelinated axons(12), is released into the cerebrospinal fluid (CSF) and serum following neuroaxonal injury(13, 14). Levels of NfL in CSF often show a strong correlation with levels in serum(15). NfL in serum has the potential to serve as a predictive biomarker for disease progression in various cerebral proteopathies, and can forecast changes in cognitive function in individuals with Alzheimer’s disease(16). An increasing amount of evidence supports the use of serum NfLfor screening and diagnosing patients with dementia(17). Serum NfL has been suggested as a valuable predictive and susceptibility biomarker for neurodegeneration in Alzheimer’s disease(18). Recent research has indicated that serum NfL levels are elevated in individuals with mild and early cognitive impairment(19, 20), suggesting its potential as a biomarker for diabetic polyneuropathy(21).

However, there are limited articles that specifically investigate the correlation between serum NfL levels and Cognitive Impairment in individuals with a diabetes diagnosis. As such, our main objective is to determine if serum NfL can serve as a potential marker for detecting early cognitive decline in diabetic patients.

## 2. Methods

### 2.1 Study population

NHANES is a large ongoing cross-sectional survey project in the US, known for its national representation and unbiased design. Conducted biennially, the project aims to track the health and nutritional status of American adults and children. Out of the total sample size of 10,175 individuals in the 2013-2014 cycle of NHANES, the study participants were limited to individuals aged 60 years and older who had been diagnosed with type 2 diabetes.Those lacking information on serum NfL, Animal Fluency exercise, education, BMI, waist circumference, smoking habits, and alcohol consumption were excluded from the study.

### 2.2 Measures

#### 2.2.1 Data Collection

Demographic variables were collected using standardized questionnaires, which included information on smoking and drinking status, gender, age, history of diabetes, marital status, education, and race.

Anthropometric data such as weight, height, and waist circumference were measured by trained health professionals. Body Mass Index (BMI) was calculated using the formula weight (kg)/height (m^2). Participants were instructed to fast for at least 8 hours before venous blood collection in the morning. Fasting blood glucose (FBG), oral glucose tolerance test (OGTT), and glycosylated hemoglobin (HbA1C) levels were measured. The OGTT involved a 75-g glucose load. Participants were classified as having type 2 diabetes if they met any of the following criteria: 1) HbA1c ≥ 6.5% or FPG ≥ 126mg/dL; 2) 2-hour plasma glucose levels ≥ 200mg/dL during OGTT; 3) self-reported physician-diagnosed diabetes; 4) self-reported use of hypoglycemic drugs (including oral medication and insulin).

#### 2.2.2 Serum Neurofilament Light Chain

The concentration of serum NfL was quantified using a highly sensitive immunoassay method utilizing acridinium ester (AE) chemiluminescence and paramagnetic particles. Analytical measurements were conducted in accordance with rigorous quality control and quality assurance protocols. The dataset obtained for analysis consisted of values falling within the range of the lower and upper detection limits.

#### 2.2.3 Cognitive function

An assessment of cognition was conducted through the Animal Fluency Score, which is a persuasive method for assessing cognitive function. This test examines executive function through categorical verbal fluency. Individuals with normal cognitive function and those with slight or more serious cognitive impairment will receive different scores(22-24). Epidemiologists frequently utilize this distinction to investigate cognitive function(23-25). During the test, participants were asked to recall as many animals as possible within a one-minute time frame. Each correct response was awarded one point. Prior to the actual test, individuals were required to complete a practice round where they were asked to name three articles of clothing. Failure to successfully complete the practice test would result in disqualification from the cognitive assessment.

### 2.3 Statistical analysis

Weighted medians with interquartile ranges were used to represent continuous variables, while the analysis was conducted using the Wilcoxon rank-sum test. For categorical variables, frequencies and weighted percentages were displayed, and the Chi-square test with the Rao and Scott second-order correction was utilized for evaluation.A multifactor analysis was conducted using weighted multivariable linear regression to examine the relationship between serum NfL and CFDAST score. Serum NfL was treated as both a categorical and continuous variable. Restricted cubic spline analysis was utilized to assess the linearity of this relationship. Data processing was carried out using R software (version 4.3.1). A statistically significant difference was defined as a p-value of less than 0.05.

## 3. Results

Serum NfL levels were analyzed and categorized into weighted tertiles. Participants with serum NfL levels below 16.8 were classified as group Q1, while those with levels above 25.7 were categorized as group Q3. Individuals with serum NfL levels between 16.8 and 25.7 were designated as group Q2. The study included 173 participants, which represents approximately 12.2 million adults in the American noninstitutionalized population. Among the participants, 51% were male, 68% were non-Hispanic white, 79% had received higher education, and the average age was 66 years old. Analysis in Table 1 revealed that participants in group Q3 exhibited higher hemoglobin A1c levels and were older compared to groups Q1 and Q2 (P < 0.01). Furthermore, significant differences in serum NfL levels were observed among the groups (Q1, Q2, and Q3; P < 0.01).

**Table 1.** Characteristics of the included participants (stratified by tertile of NfL)

| Characteristic | Overall, N = 173<br>(100%)<sup>1</sup> | NfL |  |  | P Value<sup>2</sup> |
| --- | --- | --- | --- | --- | --- |
|  |  | Q1, N = 54<br>(32%)<sup>1</sup> | Q2, N = 60<br>(34%)<sup>1</sup> | Q3, N = 59 (34%)<sup>1</sup> |  |
| Age (years) | 66.0 (63.0, 70.0) | 66.0 (63.0, 67.0) | 66.0 (61.0, 70.0) | 67.9 (64.0, 71.0) | 0.009 |
| Gender |  |  |  |  | 0.7 |
| Male | 90 (51%) | 27 (46%) | 35 (54%) | 28 (52%) |  |
| Female | 83 (49%) | 27 (54%) | 25 (46%) | 31 (48%) |  |
| Race |  |  |  |  | 0.8 |
| Hispanic and others | 72 (20%) | 23 (18%) | 24 (19%) | 25 (21%) |  |
| Non-Hispanic White | 65 (68%) | 20 (70%) | 26 (71%) | 19 (64%) |  |
| Non-Hispanic Black | 36 (12%) | 11 (12%) | 10 (9.5%) | 15 (15%) |  |
| Education |  |  |  |  | 0.7 |
| <High school | 23 (6.5%) | 10 (9.2%) | 7 (5.6%) | 6 (4.8%) |  |
| Completed High school | 32 (15%) | 7 (12%) | 12 (17%) | 13 (16%) |  |

| Characteristic | Overall, N = 173<br>(100%)<sup>1</sup> | NfL |  |  | P<br>Value<sup>2</sup> |
| --- | --- | --- | --- | --- | --- |
|  |  | Q1, N = 54<br>(32%)<sup>1</sup> | Q2, N = 60<br>(34%)<sup>1</sup> | Q3, N = 59 (34%)<sup>1</sup> |  |
| >High school | 118 (79%) | 37 (79%) | 41 (78%) | 40 (80%) |  |
| BMI | 31 (27, 36) | 30 (28, 35) | 30 (26, 35) | 33 (28, 38) | 0.4 |
| Smoke |  |  |  |  | 0.7 |
| Never | 85 (50%) | 26 (52%) | 27 (45%) | 32 (53%) |  |
| Former | 67 (41%) | 22 (37%) | 25 (48%) | 20 (39%) |  |
| Current | 21 (8.9%) | 6 (11%) | 8 (7.4%) | 7 (8.1%) |  |
| Drink | 113 (66%) | 36 (62%) | 43 (67%) | 34 (67%) | 0.8 |
| Ghb | 6.60 (6.00, 7.20) | 6.50 (5.90, 6.80) | 6.50 (6.03, 6.87) | 6.98 (6.10, 8.23) | 0.003 |
| Waist | 109 (100, 120) | 105 (100, 119) | 107 (97, 115) | 114 (104, 120) | 0.071 |
| CFDAST | 17.0 (14.0, 21.0) | 17.5 (14.0, 21.0) | 20.0 (15.3, 23.0) | 15.1(14.0, 18.0) | 0.005 |

<sup>1</sup>median (IQR) for continuous; n (%) for categorical
<sup>2</sup>Wilcoxon rank-sum test for complex survey samples; chi-squared test with Rao & Scott's second-order correction

Following adjustments for education, age, gender, GHb, and waist circumference, the weighted multivariate regression analysis model revealed a negative correlation between serum NfL (considered as a continuous variable) and CFDAST score (β = −0.05, 95% CI −0.09 to −0.02, P < 0.01; Table 2). When serum NfL was treated as a categorical variable, both group Q1 and group Q2 exhibited higher CFDAST scores compared to group Q3 after accounting for the aforementioned variables (p for trend = 0.021). Notably, Figure 2 indicated the presence of an inverted U-shaped curve linking serum NfL and CFDAST score. Specifically, when serum NfL levels were below 17 pg/mL or above 20 pg/mL, a significant negative correlation was observed. Conversely, when serum NfL levels ranged between 17 pg/mL and 20 pg/mL, the relationship was reversed (P-for-non-linear = 0.0026, P-overall < 0.05).

**Table 2.** Association between serum NfL and CFDAST score.

| serumNfL(pg/mL) | $\beta$ (95%CI) | | |
| --- | --- | --- | --- |
|  | Model 1 | Model 2 | Model 3 |
| Continuous | -0.06(-0.08– -0.03) ** | -0.05(-0.08– -0.03) ** | -0.05(-0.09– -0.02) * |
| Q3 | Ref | Ref | Ref |
| Q1 | 2.1(0.21-4.0) | 2.0(0.43-3.6) | 2.3(0.58-4.0) |
| Q2 | 3.5 (1.5–5.6) | 3.4(1.1-5.7) | 3.8(0.78-6.8) |
| *p* for trend | 0.003 | 0.008 | 0.021 |

Model 1, Unadjusted;
Model 2, adjusted for Age and Gender;
Model 3, adjusted for Age,Gender,Education,Ghb,Waist;
\*0.001 < *P* < 0.01, \*\**P* < 0.001

**Fig. 1.**
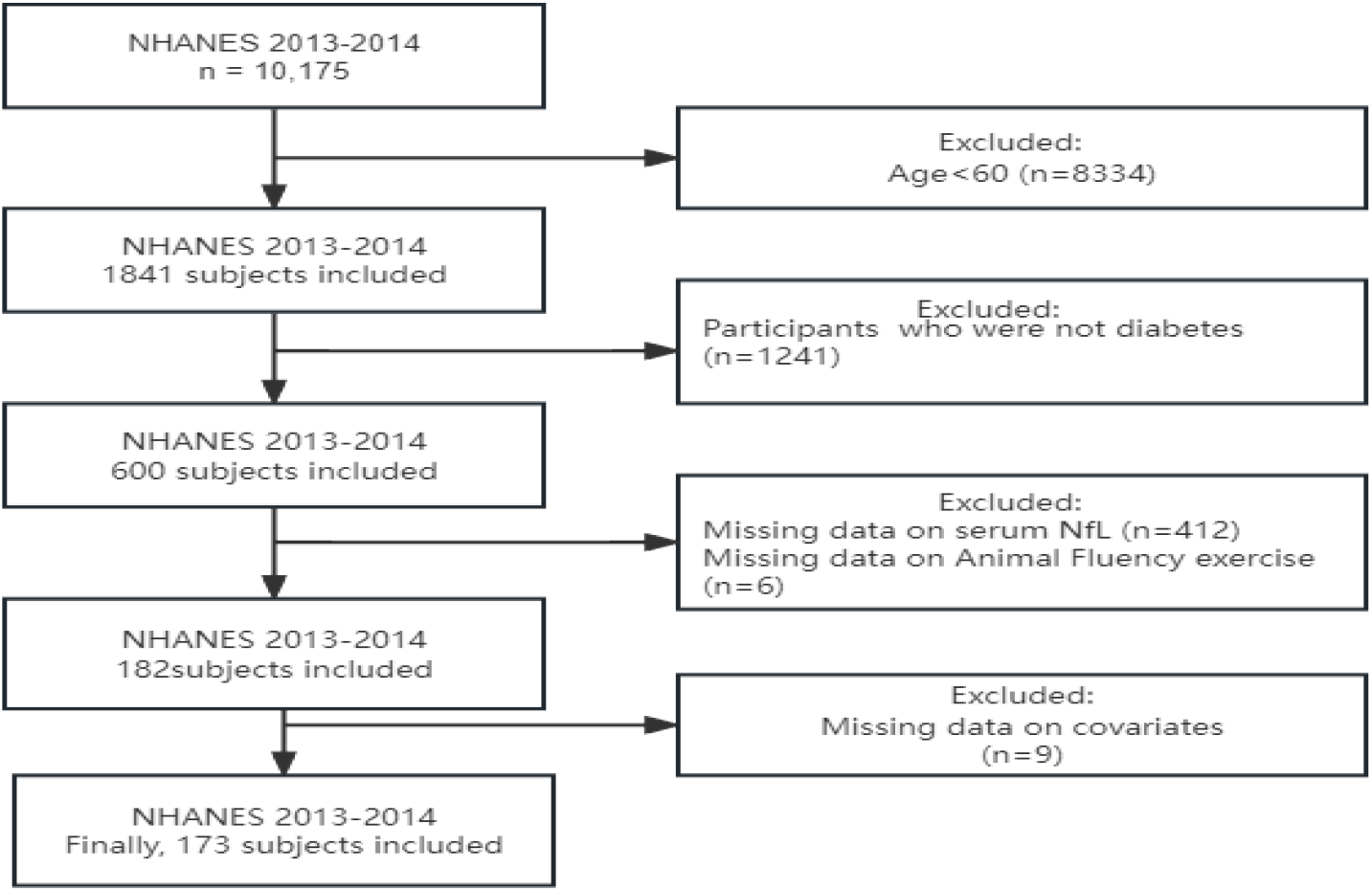
Flow chat for the study design and participants.

**Fig. 2.**
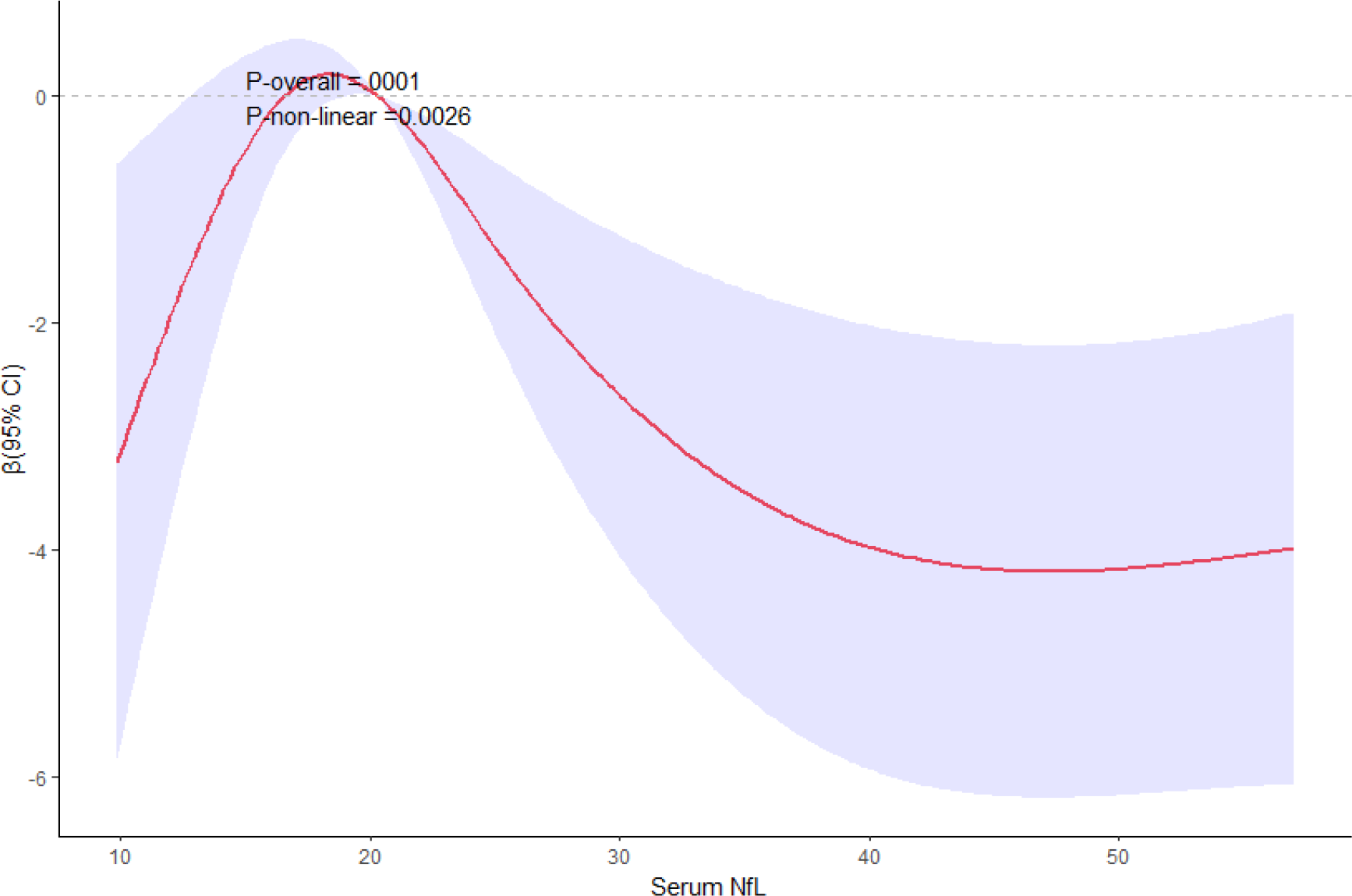
Restricted cubic spline plot of the relationship between serum NfL and CFDAST score. Adjustment was made for Age,Gender,Education,Ghb and Waist.Data were weighted during analysis

## 4. Discussion

Serum NfL levels were found to be associated with cognitive function in participants with diabetes. Additionally, we were the first to demonstrate a non-linear, inverted U-shaped relationship between them. This phenomenon may be linked to the body’s compensatory mechanism. We hypothesize that when serum NfL levels fall within a specific range, the body is able to maintain a balance between neuronal death and survival, thereby supporting cognitive function.

This study aligns with the research conducted by Noriko et al.(26), which suggests that serum NfL can be used to screen for early cognitive decline in patients with diabetes. However, their model did not account for education level as a covariate in the analysis,and the patients included in their study were exclusively from outpatient clinics, potentially introducing selection bias. Moreover, cognitive decline and dementia are most common in individuals over the age of 60 in the general population(27, 28). Therefore, screening for early cognitive decline in general population with diabetes holds significant importance. While the exact reasons for the increased risk of cognitive disorders in people with diabetes remain unclear, it is likely due to a combination of factors with considerable inter-individual variation(3). The primary pathological mechanisms of dementia involve tau protein accumulation leading to neuronal loss and vascular brain damage(29). In addition to elevating the risk of stroke and cerebral small vessel disease(30), diabetes can also heighten the risk of neurodegeneration through tau-mediated pathways(1, 31, 32). Studies have shown that serum NfL can differentiate between neurodegenerative dementias and healthy individuals(17). Furthermore, the levels of NfL in the blood have been linked to brain structures associated with small vessel vascular disease(33), and its correlation with white matter hyperintensity volume may partially explain its connection to dementia events related to cerebral small vessel disease(34). Recent research indicates that higher levels of serum NfL are related to lower total brain volume and the presence of infarcts(35). Taking all these findings into consideration, it is reasonable to conclude that serum NfL is linked to cognitive functioning in individuals diagnosed with type 2 diabetes.

Our research has both strengths and weaknesses. We were the first to establish a connection between serum NfL and cognitive functioning in the general population with diabetes. The 173 individuals we studied can be considered representative of approximately 12 million civilian, non-institutionalized older adults in the United States. Furthermore, we were the first to identify a non-linear relationship between serum NfL and cognitive levels in elderly diabetic patients. However, our study is limited by its reliance on a cross-sectional database, which prevents us from establishing causation. Additionally, potential confounders such as physical activity may have influenced our results. Lastly, we focused solely on one domain of cognitive function, potentially limiting the generalizability of our findings to other domains.

## 5. Conclusion

Previous research(36) has identified the serum NfL reference limit for individuals over 60 years of age as 20pg/ml. Analysis of Figure 2 reveals a negative correlation between serum NfL levels exceeding this reference limit and cognitive scores. Therefore, our study proposes that serum NfL could potentially serve as a valuable biomarker for detecting early cognitive decline in elderly diabetic patients.

## Data Availability

All data files are available from the NHANES database

https://www.cdc.gov/nchs/nhanes/index.htm?CDC_AA_refVal=https%3A%2F%2Fwww.cdc.gov%2Fnchs%2Fnhanes.htm

## Funding

This research did not receive any specific grant from funding agencies in the public, commercial, or not-for-profit sectors.

## CRediT authorship contribution statement

**Li Li:** Conceptualization, Data curation, Methodology, Software, Writing – original draft. **Cheng-Bo Li:** Formal analysis, Software, Writing – original draft. **Li Zhu:** Software, Validation, Writing–original draft. **Jiang Zhu:** Visualization, Data curation, Software Supervision, Validation.

## Declaration of Competing Interest

No potential conflicts of interest relevant to this article were reported.

## Acknowledgments

The authors thank all of the survey teams and all of the study participants in the NHANES for their contributions and dedication to this study.

## Notes

### Competing Interest Statement

The authors have declared no competing interest.

### Funding Statement

The author(s) received no specific funding for this work.

### Author Declarations

You can find related information from website:https://www.cdc.gov/nchs/nhanes/irba98.htm and website: https://www.cdc.gov/nchs/nhanes/genetics/genetic_participants.htm.

## References

1. Srikanth V, Sinclair AJ, Hill-Briggs F, Moran C, Biessels GJ. Type 2 diabetes and cognitive dysfunction-towards effective management of both comorbidities.Lancet Diabetes Endocrinol. 1st June, 2020.10.1016/s2213-8587(20)30118-22020

2. Bouchard MF, Oulhote Y, Sagiv SK, Saint-Amour D, Weuve J. Polychlorinated biphenyl exposures and cognition in older U.S. adults: NHANES (1999-2002).Environ Health Perspect. 15th January, 2014.10.1289/ehp.13065322014

3. Biessels GJ, Despa F. Cognitive decline and dementia in diabetes mellitus: mechanisms and clinical implications.Nat Rev Endocrinol. 15th October, 2018.10.1038/s41574-018-0048-72018

4. Koekkoek PS, Kappelle LJ, van den Berg E, Rutten GE, Biessels GJ. Cognitive function in patients with diabetes mellitus: guidance for daily care.Lancet Neurol. 1st March, 2015.10.1016/s1474-4422(14)70249-22015

5. Chen Q, Zhu S, Shang J, Fang Q, Xue Q, Hua J. Trends in Cognitive Function Before and After Diabetes Onset: The China Health and Retirement Longitudinal Study.Neurology. 9th April, 2024.10.1212/WNL.00000000002091652024

6. Monette MC, Baird A, Jackson DL. A meta-analysis of cognitive functioning in nondemented adults with type 2 diabetes mellitus.Can J Diabetes. 1st December, 2014.10.1016/j.jcjd.2014.01.0142014

7. Biessels GJ, Strachan MW, Visseren FL, Kappelle LJ, Whitmer RA. Dementia and cognitive decline in type 2 diabetes and prediabetic stages: towards targeted interventions.Lancet Diabetes Endocrinol. 1st March, 2014.10.1016/s2213-8587(13)70088-32014

8. Pappas C, Andel R, Infurna FJ, Seetharaman S. Glycated haemoglobin (HbA1c), diabetes and trajectories of change in episodic memory performance.J Epidemiol Community Health. 1st February, 2017.10.1136/jech-2016-2075882017

9. Bangen KJ, Gu Y, Gross AL, Schneider BC, Skinner JC, Benitez A, et al. Relationship Between Type 2 Diabetes Mellitus and Cognitive Change in a Multiethnic Elderly Cohort.J Am Geriatr Soc. 1st June, 2015.10.1111/jgs.134412015

10. Yaffe K, Falvey C, Hamilton N, Schwartz AV, Simonsick EM, Satterfield S, et al. Diabetes, glucose control, and 9-year cognitive decline among older adults without dementia.Arch Neurol. 15th September, 2012.10.1001/archneurol.2012.11172012

11. Ehtewish H, Arredouani A, El-Agnaf O. Diagnostic, Prognostic, and Mechanistic Biomarkers of Diabetes Mellitus-Associated Cognitive Decline.Int J Mol Sci. 30th May 2022.10.3390/ijms231161442022

12. Quiroz YT, Zetterberg H, Reiman E.M, Chen Y, Su Y, Fox-Fuller JT, et al. Plasma neurofilament light chain in the presenilin 1 E280A autosomal dominant Alzheimer’s disease kindred: a cross-sectional and longitudinal cohort study.Lancet Neurol. Jun, 2020.10.1016/S1474-4422(20)30137-X2020

13. Siller N, Kuhle J, Muthuraman M, Barro C, Uphaus T, Groppa S, et al. Serum neurofilament light chain is a biomarker of acute and chronic neuronal damage in early multiple sclerosis.Mult Scler. April, 2019.10.1177/13524585187656662019

14. Ziemssen T, Arnold D L, Alvarez E, Cross A.H, Willi R, Li B, et al. Prognostic Value of Serum Neurofilament Light Chain for Disease Activity and Worsening in Patients With Relapsing Multiple Sclerosis: Results From the Phase 3 ASCLEPIOS I and II Trials.Front Immunol. 2022.10.3389/fimmu.2022.8525632022

15. Desai P, Dhana K, DeCarli C, Wilson RS, McAninch EA, Evans DA, et al. Examination of Neurofilament Light Chain Serum Concentrations, Physical Activity, and Cognitive Decline in Older Adults.JAMA Netw Open. 1st March, 2022.10.1001/jamanetworkopen.2022.35962022

16. Preische O, Schultz S.A, Apel A, Kuhle J, Kaeser SA, Barro C, et al. Serum neurofilament dynamics predicts neurodegeneration and clinical progression in presymptomatic Alzheimer’s disease.Nat Med. February, 2019.10.1038/s41591-018-0304-32019

17. Baiardi S, Quadalti C, Mammana A, Dellavalle S, Zenesini C, Sambati L, et al. Diagnostic value of plasma p-tau181, NfL, and GFAP in a clinical setting cohort of prevalent neurodegenerative dementias.Alzheimers Res Ther. 12th October, 2022.10.1186/s13195-022-01093-62022

18. Jung Y, Damoiseaux JS. The potential of blood neurofilament light as a marker of neurodegeneration for Alzheimer’s disease.Brain. 4th January 2024.10.1093/brain/awad2672024

19. Gaur A, Rivet L, Mah E, Bawa KK, Gallagher D, Herrmann N, et al. Novel fluid biomarkers for mild cognitive impairment: A systematic review and meta-analysis.Ageing Res Rev. November, 2023.10.1016/j.arr.2023.1020462023

20. De Meyer S, Blujdea ER, Schaeverbeke J, Reinartz M, Luckett ES, Adamczuk K, et al. Longitudinal associations of serum biomarkers with early cognitive, amyloid and grey matter changes.Brain. 1st March, 2024.10.1093/brain/awad3302024

21. Maatta LL, Andersen ST, Parkner T, Hviid CVB, Bjerg L, Kural MA, et al. Longitudinal Change in Serum Neurofilament Light Chain in Type 2 Diabetes and Early Diabetic Polyneuropathy: ADDITION-Denmark.Diabetes Care. 19th Marth, 2024.10.2337/dc23-22082024

22. Henry JD, Crawford JR, Phillips LH. Verbal fluency performance in dementia of the Alzheimer’s type: a meta-analysis.Neuropsychologia. 15th June, 2004.10.1016/j.neuropsychologia.2004.02.0012004

23. Clark LJ, Gatz M, Zheng L, Chen YL, McCleary C, Mack WJ. Longitudinal verbal fluency in normal aging, preclinical, and prevalent Alzheimer’s disease.Am J Alzheimers Dis Other Demen. 15th December, 2009.10.1177/15333175093451542009

24. Canning SJ, Leach L, Stuss D, Ngo L, Black SE. Diagnostic utility of abbreviated fluency measures in Alzheimer disease and vascular dementia.Neurology. 24th February, 2004.10.1212/wnl.62.4.5562004

25. Grundman M, Petersen RC, Ferris S.H, Thomas RG, Aisen PS, Bennett DA, et al. Mild cognitive impairment can be distinguished from Alzheimer disease and normal aging for clinical trials.Arch Neurol. Jan, 2004.10.1001/archneur.61.1.592004

26. Marutani N, Akamine S, Kanayama D, Gotoh S, Yanagida K, Maruyama R, et al. Plasma NfL is associated with mild cognitive decline in patients with diabetes.Psychogeriatrics. May, 2022.10.1111/psyg.128192022

27. Sachdev PS, Lipnicki DM, Kochan NA, Crawford JD, Thalamuthu A, Andrews G, et al. The Prevalence of Mild Cognitive Impairment in Diverse Geographical and Ethnocultural Regions: The COSMIC Collaboration.PLoS One. 15th June, 2015.10.1371/journal.pone.01423882015

28. Saeedi P, Petersohn I, Salpea P, Malanda B, Karuranga S, Unwin N, et al. Global and regional diabetes prevalence estimates for 2019 and projections for 2030 and 2045: Results from the International Diabetes Federation Diabetes Atlas, 9 th edition.Diabetes Res Clin Pract. 15th November, 2019.10.1016/j.diabres.2019.1078432019

29. Geijselaers SLC, Sep SJS, Stehouwer CDA, Biessels GJ. Glucose regulation, cognition, and brain MRI in type 2 diabetes: a systematic review.Lancet Diabetes Endocrinol. 1st January, 2015.10.1016/s2213-8587(14)70148-22015

30. Abner EL, Nelson PT, Kryscio RJ, Schmitt FA, Fardo DW, Woltjer RL, et al. Diabetes is associated with cerebrovascular but not Alzheimer’s disease neuropathology.Alzheimers Dement. August, 2016.10.1016/j.jalz.2015.12.0062016

31. Moran C, Phan TG, Chen J, Blizzard L, Beare R, Venn A, et al. Brain atrophy in type 2 diabetes: regional distribution and influence on cognition.Diabetes Care. December, 2013.10.2337/dc13-01432013

32. Moran C, Beare R, Phan TG, Bruce DG, Callisaya ML, Srikanth V, et al. Type 2 diabetes mellitus and biomarkers of neurodegeneration.Neurology. 29th September, 2015.10.1212/WNL.00000000000019822015

33. Rübsamen N, Maceski A, Leppert D, Benkert P, Kuhle J, Wiendl H, et al. Serum neurofilament light and tau as prognostic markers for all-cause mortality in the elderly general population-an analysis from the MEMO study.BMC Med. 15th February, 2021.10.1186/s12916-021-01915-82021

34. van Gennip ACE, Satizabal CL, Tracy RP, Sigurdsson S, Gudnason V, Launer LJ, et al. Associations of plasma NfL, GFAP, and t-tau with cerebral small vessel disease and incident dementia: longitudinal data of the AGES-Reykjavik Study.Geroscience. 15th February, 2024.10.1007/s11357-023-00888-12024

35. Twait E. L, Gerritsen L, Moonen JEF, Verberk IMW, Teunissen CE, Visser PJ, et al. Plasma Markers of Alzheimer’s Disease Pathology, Neuronal Injury, and Astrocytic Activation and MRI Load of Vascular Pathology and Neurodegeneration: The SMART-MR Study.J Am Heart Assoc. 20th February, 2024.10.1161/jaha.123.0321342024

36. Simren J, Andreasson U, Gobom J, Suarez Calvet M, Borroni B, Gillberg C, et al. Establishment of reference values for plasma neurofilament light based on healthy individuals aged 5-90 years.Brain Commun. 2022.10.1093/braincomms/fcac1742022

